# Quality-adjusted life expectancy norms for the English population

**DOI:** 10.1101/2021.12.13.21267671

**Authors:** Paul Schneider, Simon McNamara, James Love-Koh, Tim Doran, Nils Gutacker

## Abstract

**Objective:** The National Institute for Health and Care Excellence in England has proposed severity-of-disease modifiers that give greater weight to health benefits accruing to patients who experience a larger shortfall in quality-adjusted life years (QALYs) under current standard of care compared to healthy individuals. This requires an estimate of quality-adjusted life expectancy (QALE) of the general population by age and sex. Previous QALE population norms are based on nearly 30-year old assessments of HRQoL in the general population. This study provides updated QALE estimates for the English population by age and sex.

**Methods:** EQ-5D-5L data for 14,412 participants from the Health Survey for England (waves 2017 and 2018) were pooled and HRQoL population norms were calculated. These norms were combined with official life tables from the Office for National Statistics for 2017-2019 using the Sullivan method to derive QALE estimates by age and sex. Values were discounted using 0%, 1.5% and 3.5% discount rates.

**Results:** QALE at birth is 68.04 QALYs for men and 68.48 QALYs for women. These values are lower than previously published QALE population norms based on older HRQoL data. Additional data tables and figures are made available through an interactive web application: https://r4scharr.shinyapps.io/shortfall/.

**Conclusions:** This study provides new QALE population norms for England that serve to establish absolute and relative QALY shortfalls for the purpose of health technology assessments.

## Introduction

The National Institute for Health and Care Excellence (NICE) has recently published proposals for revised methods of health technology evaluation (Catchpole and Barrett 2020, NICE 2021). As part of these revisions, NICE proposes to introduce a new severity-of-disease modifier: a mechanism designed to enable their advisory committees to formally, quantitatively, and transparently grant additional weight to incremental quality-adjusted life-years (QALYs) provided to people with more severe health conditions. This mechanism - if implemented - would further the development introduced through the end-of-life criterion in 2009, in which NICE moved away from the long-held view that QALY gains are equally valuable independent of who they accrue to (NICE 2009).

For the purpose of this modifier, NICE propose to define the severity of a health condition using two metrics: absolute QALY-shortfall and proportional QALY-shortfall. Absolute shortfall is quantified as the absolute number of future QALYs an individual can expect to lose as a result of that condition, given currently available interventions (Arnberg 2012). In contrast, proportional shortfall is quantified as the proportion of future QALYs a person can expect to lose as a result of their condition (Stolk et al. 2004). If the magnitude of these shortfall metrics is sufficiently large, any incremental gains in QALYs achieved by a new health technology would be assigned weights greater than one, thus increasing the effective cost-effectiveness threshold for these interventions. At present, absolute shortfall is used as severity modifier in Norway (Ottersen et al. 2016, Statens legemiddelverk 2020), whilst proportional shortfall is applied in the Netherlands (Reckers-Droog et al. 2018, Zorginstituut Nederland 2015).

Both shortfall metrics require two pieces of information: (i) an estimate of the number of future QALYs people who receive current standard of care can expect to experience in their lifetime, and (ii) an estimate of the number of future QALYs that individuals with the condition would have experienced had they been healthy. The first of these is a standard output of a cost-utility analysis, and so is already available as part of a NICE appraisal conducted using current methods. The second is not routinely calculated as part of a current NICE assessment, but will be required if NICE’s proposed methods are implemented. In practical terms, this information could be estimated independently by each of the stakeholders making submissions to NICE, or appraising evidence on behalf of NICE. Alternatively, reference values (‘population norms’) could be established outside the review process in order to improve consistency across appraisals, reduce the burden on stakeholders, and provide a basis for appraisal-specific modification if warranted.

Quality-adjusted life-expectancy (QALE) population norms for several countries including England have been published by Heijink et al. (2011). The authors combined mortality data from the Human Mortality Database (2013) and EQ-5D-3L data from the 1993 Measuring and Valuing Health (MVH Group 1995) study via a life table approach to derive estimates of QALE stratified by age and sex. More recently, Palmer et al. (2021) used a two-state Markov model to provide updated QALE estimates based on UK population life tables for the period 2017 to 2019. Both studies rely on the rather dated EQ-5D population norms from the MVH study and focus on the -3L version of the instrument despite the EQ-5D-5L instrument - a newer version of the instrument with a more detailed descriptive system (Herdman et al. 2011) - being rapidly adopted in clinical trials and observational studies. Furthermore, the EQ-5D-3L population norms were based on relatively small samples of individuals, which induces considerable sampling uncertainty in any assessment of severity of disease (Kind et al. 1999). This limits the usefulness of existing QALE population norms for NICE decision-making.

In this paper, we report more recent (2017/18) estimates of the QALE of the English population, drawing on a large dataset of quality of life measurements collected using the EQ-5D-5L instrument. In parallel, we publish an R Shiny online tool inspired by the iDBC platform of Versteegh et al. (2019). Our tool enables users to combine QALE population norms with the outputs of an economic model to estimate the absolute and proportional QALY-shortfall associated with a condition. It is available at: https://r4scharr.shinyapps.io/shortfall/

## Methods

To derive QALE population norms, we combined age- and sex-specific EQ-5D-5L utility scores with national life tables of the English population.

National life tables (pooled for 2017-2019) were taken from the Office of National Statistics (2021). We used the Chiang II method to derive crude age- and sex-specific life expectancies (Chiang 1984). The life expectancy at the start of age interval i is accordingly estimated by dividing the number of years lived in that and all successive intervals by the number of people alive at the beginning of interval i.

Information on self-reported EQ-5D-5L health state profiles were retrieved from the 2017 and 2018 waves of the Health Survey for England, which is a long-running survey of the English population (NatCen 2019, 2020). We used the individual sampling weights to adjust the sample for non-reponders, and to make it nationally representative in terms of age, sex and geography.

Health states were valued in terms of utilities using the van Hout et al. (2012) crosswalk method, which is the approach currently recommended by NICE. This method maps the EQ-5D-5L health states to the EQ-5D-3L value set for the UK. To derive QALEs, mean utility scores by age and sex were calculated, and then combined with life expectancy estimates, using the Sullivan method (Sullivan, 1971). Age- and sex-specific QALE estimates for the English population are reported for a 1.5% and a 3.5% annual discount rate, as well as un-discounted.

For the analysis, we had to make several assumptions. Firstly, while life tables are reported by single year of age, EQ-5D-5L data were only available according to the grouped age variable in the Health Survey for England (16-17, 18-9, followed by 5-year bands, up until 90+). We assumed that HRQoL was constant within each age band. Secondly, the HSE does not contain any EQ-5D-5L data for children under the age of 16. It was assumed that children aged 0-15 had the same HRQoL as those aged 16-17. Thirdly, the calculation of life expectancies was based on the assumption that individuals dying at a given year of age had an average survival of six months (half-cycle correction). The life table was closed at a maximum age of 100.

To help stakeholders estimate the absolute and proportional QALY shortfall, we developed an interactive web application (https://r4scharr.shinyapps.io/shortfall/). The ‘QALY Shortfall Calculator’ can be used to compute the difference between the QALE of individuals without and individuals with a particular disease (the estimate for the latter obviously needs to be supplied by the user). The app allows the user to adjust the age and male/female distribution of the patient group, as well as the discount rate that is applied.

The R source code of the web application, as well as the code used to generate the results reported in this paper, is available online at: https://github.com/bitowaqr/shortfall.

## Results

A total of 16,175 individuals participated in the HSE in 2017 (n=7,997) and 2018 (n=8,178). Of these, 1,762 (10.9%) respondents did not report their EQ-5D-5L health state and were excluded from the analysis. This left 14,413 individuals for the estimation of utility scores by age and sex.

Table 1 shows average EQ-5D-5L utility scores (based on the van Hout et al. crosswalk method) by age for male and female respondents.

**Table 1.** Mean EQ-5D-5L utility scores */* HRQoL by age group and sex and 95% confidence intervalls (based on bootstrapping with 10,000 iterations).

| Age | Female |  | Male |  |
| --- | --- | --- | --- | --- |
|  | Mean (95%CI) | n | Mean (95%CI) | n |
| 16-17 | 0.881 (0.874; 0.904) | 151 | 0.916 (0.909; 0.936) | 146 |
| 18-19 | 0.864 (0.852; 0.899) | 122 | 0.933 (0.928; 0.951) | 110 |
| 20-24 | 0.866 (0.860; 0.885) | 370 | 0.895 (0.890; 0.912) | 298 |
| 25-29 | 0.873 (0.868; 0.889) | 515 | 0.895 (0.890; 0.913) | 366 |
| 30-34 | 0.870 (0.865; 0.885) | 669 | 0.916 (0.912; 0.928) | 450 |
| 35-39 | 0.857 (0.853; 0.871) | 722 | 0.862 (0.855; 0.886) | 465 |
| 40-44 | 0.850 (0.845; 0.867) | 668 | 0.870 (0.865; 0.888) | 498 |
| 45-49 | 0.815 (0.810; 0.833) | 693 | 0.824 (0.816; 0.847) | 527 |
| 50-54 | 0.805 (0.799; 0.824) | 729 | 0.837 (0.832; 0.854) | 529 |
| 55-59 | 0.802 (0.796; 0.821) | 730 | 0.817 (0.811; 0.836) | 582 |
| 60-64 | 0.784 (0.778; 0.805) | 608 | 0.809 (0.803; 0.830) | 532 |
| 65-69 | 0.782 (0.776; 0.802) | 619 | 0.798 (0.792; 0.818) | 568 |
| 70-74 | 0.787 (0.782; 0.804) | 619 | 0.802 (0.797; 0.821) | 505 |
| 75-79 | 0.741 (0.734; 0.764) | 399 | 0.791 (0.784; 0.813) | 335 |
| 80-84 | 0.717 (0.709; 0.745) | 268 | 0.773 (0.764; 0.802) | 233 |
| 85-89 | 0.665 (0.653; 0.706) | 145 | 0.718 (0.703; 0.765) | 126 |
| 90+ | 0.665 (0.650; 0.716) | 67 | 0.663 (0.642; 0.728) | 49 |

Table 2 shows the age- and sex-specific period life expectancy (LE) and QALE, undiscounted and with a 1.5% and 3.5% discount rate applied. Women are expected to live considerably longer (+3.7 years) but have similar undiscounted QALE at birth as men (+0.4 QALYs).

**Table 2.** QALE by age and sex with 0%, 1.5% and 3.5% discount rates (based on van Hout et al. 2012 cross walk utilities)

| Age | Female |  |  |  | Male |  |  |  |
| --- | --- | --- | --- | --- | --- | --- | --- | --- |
|  | LE | QALE |  |  | LE | QALE |  |  |
|  |  | 0 % | 1.5 % | 3.5 % |  | 0 % | 1.5 % | 3.5 % |
| 0 | 83.33 | 68.68 | 40.15 | 23.73 | 79.67 | 68.28 | 40.63 | 24.35 |
| 1 | 82.63 | 68.04 | 40.01 | 23.74 | 79.02 | 67.65 | 40.49 | 24.36 |
| 2 | 81.65 | 67.18 | 39.72 | 23.66 | 78.04 | 66.75 | 40.17 | 24.27 |
| 3 | 80.66 | 66.30 | 39.43 | 23.58 | 77.05 | 65.85 | 39.85 | 24.17 |
| 4 | 79.67 | 65.43 | 39.13 | 23.50 | 76.06 | 64.94 | 39.52 | 24.07 |
| 5 | 78.67 | 64.55 | 38.83 | 23.41 | 75.06 | 64.03 | 39.19 | 23.97 |
| 6 | 77.68 | 63.68 | 38.52 | 23.32 | 74.07 | 63.12 | 38.85 | 23.86 |
| 7 | 76.68 | 62.80 | 38.20 | 23.22 | 73.08 | 62.21 | 38.51 | 23.75 |
| 8 | 75.69 | 61.92 | 37.89 | 23.13 | 72.08 | 61.30 | 38.16 | 23.64 |
| 9 | 74.69 | 61.05 | 37.56 | 23.03 | 71.09 | 60.39 | 37.81 | 23.52 |
| 10 | 73.70 | 60.17 | 37.23 | 22.92 | 70.09 | 59.47 | 37.45 | 23.40 |
| 11 | 72.70 | 59.29 | 36.90 | 22.81 | 69.09 | 58.56 | 37.08 | 23.27 |
| 12 | 71.71 | 58.42 | 36.56 | 22.70 | 68.10 | 57.65 | 36.71 | 23.14 |
| 13 | 70.71 | 57.54 | 36.22 | 22.59 | 67.11 | 56.74 | 36.34 | 23.00 |
| 14 | 69.72 | 56.66 | 35.87 | 22.47 | 66.12 | 55.83 | 35.96 | 22.86 |
| 15 | 68.72 | 55.79 | 35.52 | 22.34 | 65.12 | 54.92 | 35.57 | 22.72 |
| 16 | 67.73 | 54.91 | 35.16 | 22.22 | 64.13 | 54.02 | 35.18 | 22.57 |
| 17 | 66.74 | 54.04 | 34.80 | 22.09 | 63.15 | 53.11 | 34.79 | 22.42 |
| 18 | 65.75 | 53.17 | 34.43 | 21.95 | 62.17 | 52.21 | 34.39 | 22.26 |
| 19 | 64.76 | 52.31 | 34.08 | 21.83 | 61.19 | 51.30 | 33.97 | 22.08 |
| 20 | 63.78 | 51.46 | 33.72 | 21.70 | 60.22 | 50.39 | 33.55 | 21.90 |
| 21 | 62.79 | 50.60 | 33.35 | 21.57 | 59.25 | 49.52 | 33.16 | 21.75 |
| 22 | 61.80 | 49.75 | 32.98 | 21.43 | 58.28 | 48.65 | 32.77 | 21.60 |
| 23 | 60.81 | 48.89 | 32.60 | 21.29 | 57.30 | 47.78 | 32.36 | 21.44 |
| 24 | 59.82 | 48.04 | 32.22 | 21.15 | 56.33 | 46.90 | 31.96 | 21.27 |
| 25 | 58.84 | 47.18 | 31.83 | 20.99 | 55.36 | 46.03 | 31.54 | 21.10 |
| 26 | 57.85 | 46.32 | 31.43 | 20.83 | 54.39 | 45.16 | 31.13 | 20.92 |
| 27 | 56.87 | 45.46 | 31.03 | 20.66 | 53.42 | 44.29 | 30.70 | 20.74 |
| 28 | 55.88 | 44.60 | 30.61 | 20.49 | 52.45 | 43.42 | 30.27 | 20.55 |
| 29 | 54.90 | 43.74 | 30.20 | 20.31 | 51.48 | 42.55 | 29.84 | 20.36 |
| 30 | 53.92 | 42.88 | 29.77 | 20.12 | 50.51 | 41.69 | 29.40 | 20.16 |
| 31 | 52.93 | 42.02 | 29.35 | 19.93 | 49.55 | 40.80 | 28.93 | 19.93 |
| 32 | 51.95 | 41.17 | 28.91 | 19.74 | 48.59 | 39.92 | 28.45 | 19.70 |
| 33 | 50.98 | 40.32 | 28.48 | 19.54 | 47.62 | 39.03 | 27.97 | 19.45 |
| 34 | 50.00 | 39.46 | 28.03 | 19.33 | 46.66 | 38.15 | 27.49 | 19.20 |
| 35 | 49.03 | 38.61 | 27.59 | 19.11 | 45.71 | 37.27 | 27.00 | 18.95 |
| 36 | 48.05 | 37.78 | 27.15 | 18.91 | 44.75 | 36.44 | 26.55 | 18.74 |
| 37 | 47.08 | 36.94 | 26.70 | 18.69 | 43.80 | 35.62 | 26.10 | 18.52 |
| 38 | 46.11 | 36.11 | 26.25 | 18.47 | 42.85 | 34.80 | 25.65 | 18.30 |
| 39 | 45.15 | 35.28 | 25.79 | 18.25 | 41.90 | 33.98 | 25.19 | 18.07 |
| 40 | 44.18 | 34.45 | 25.33 | 18.01 | 40.95 | 33.16 | 24.73 | 17.83 |
| 41 | 43.22 | 33.63 | 24.87 | 17.78 | 40.01 | 32.33 | 24.25 | 17.58 |
| 42 | 42.26 | 32.81 | 24.40 | 17.54 | 39.07 | 31.51 | 23.77 | 17.32 |
| 43 | 41.30 | 31.99 | 23.93 | 17.29 | 38.14 | 30.69 | 23.28 | 17.06 |
| 44 | 40.34 | 31.18 | 23.45 | 17.04 | 37.21 | 29.88 | 22.79 | 16.79 |
| 45 | 39.39 | 30.37 | 22.97 | 16.77 | 36.28 | 29.07 | 22.29 | 16.51 |
| 46 | 38.45 | 29.59 | 22.52 | 16.54 | 35.36 | 28.31 | 21.84 | 16.27 |
| 47 | 37.50 | 28.82 | 22.06 | 16.30 | 34.44 | 27.55 | 21.38 | 16.03 |
| 48 | 36.56 | 28.05 | 21.60 | 16.05 | 33.53 | 26.79 | 20.92 | 15.78 |
| 49 | 35.63 | 27.29 | 21.14 | 15.80 | 32.62 | 26.04 | 20.46 | 15.52 |

LE = life expectancy; QALE = quality-adjusted life expectancy

**Table 2. (continued) QALE by age and sex with 0%, 1.5% and 3.5% discount rates (based on van Hout et al. 2012 cross walk utilities)**
| Age | Female |  |  |  | Male |  |  |  |
| --- | --- | --- | --- | --- | --- | --- | --- | --- |
|  | LE |  | QALE |  | LE |  | QALE |  |
|  |  | 0 % | 1.5 % | 3.5 % |  | 0 % | 1.5 % | 3.5 % |
| 50 | 34.69 | 26.52 | 20.67 | 15.54 | 31.71 | 25.30 | 19.99 | 15.26 |
| 51 | 33.77 | 25.77 | 20.20 | 15.28 | 30.81 | 24.54 | 19.50 | 14.97 |
| 52 | 32.84 | 25.03 | 19.73 | 15.02 | 29.92 | 23.79 | 19.02 | 14.69 |
| 53 | 31.92 | 24.28 | 19.26 | 14.75 | 29.03 | 23.04 | 18.52 | 14.39 |
| 54 | 31.00 | 23.54 | 18.78 | 14.47 | 28.15 | 22.29 | 18.02 | 14.08 |
| 55 | 30.09 | 22.80 | 18.30 | 14.19 | 27.27 | 21.55 | 17.52 | 13.77 |
| 56 | 29.18 | 22.07 | 17.82 | 13.90 | 26.39 | 20.83 | 17.04 | 13.47 |
| 57 | 28.28 | 21.34 | 17.33 | 13.60 | 25.53 | 20.12 | 16.55 | 13.17 |
| 58 | 27.38 | 20.61 | 16.84 | 13.30 | 24.67 | 19.42 | 16.06 | 12.86 |
| 59 | 26.49 | 19.89 | 16.35 | 12.99 | 23.82 | 18.72 | 15.58 | 12.55 |
| 60 | 25.61 | 19.18 | 15.85 | 12.67 | 22.98 | 18.03 | 15.08 | 12.22 |
| 61 | 24.73 | 18.48 | 15.37 | 12.36 | 22.14 | 17.35 | 14.60 | 11.90 |
| 62 | 23.86 | 17.80 | 14.88 | 12.05 | 21.32 | 16.68 | 14.11 | 11.58 |
| 63 | 23.00 | 17.12 | 14.40 | 11.73 | 20.51 | 16.01 | 13.63 | 11.25 |
| 64 | 22.15 | 16.44 | 13.91 | 11.41 | 19.71 | 15.36 | 13.15 | 10.92 |
| 65 | 21.30 | 15.77 | 13.42 | 11.08 | 18.91 | 14.71 | 12.66 | 10.58 |
| 66 | 20.46 | 15.11 | 12.93 | 10.74 | 18.13 | 14.08 | 12.19 | 10.25 |
| 67 | 19.62 | 14.45 | 12.43 | 10.40 | 17.36 | 13.47 | 11.72 | 9.92 |
| 68 | 18.80 | 13.79 | 11.94 | 10.04 | 16.60 | 12.85 | 11.25 | 9.58 |
| 69 | 17.98 | 13.15 | 11.44 | 9.69 | 15.86 | 12.25 | 10.78 | 9.24 |
| 70 | 17.17 | 12.50 | 10.94 | 9.32 | 15.12 | 11.65 | 10.31 | 8.89 |
| 71 | 16.38 | 11.86 | 10.44 | 8.95 | 14.38 | 11.05 | 9.83 | 8.53 |
| 72 | 15.58 | 11.23 | 9.93 | 8.56 | 13.66 | 10.47 | 9.36 | 8.17 |
| 73 | 14.81 | 10.60 | 9.42 | 8.17 | 12.96 | 9.89 | 8.88 | 7.80 |
| 74 | 14.06 | 9.99 | 8.92 | 7.78 | 12.27 | 9.33 | 8.42 | 7.44 |
| 75 | 13.31 | 9.38 | 8.42 | 7.38 | 11.60 | 8.78 | 7.96 | 7.07 |
| 76 | 12.58 | 8.83 | 7.97 | 7.03 | 10.95 | 8.25 | 7.52 | 6.72 |
| 77 | 11.87 | 8.30 | 7.52 | 6.67 | 10.32 | 7.74 | 7.09 | 6.37 |
| 78 | 11.18 | 7.77 | 7.08 | 6.32 | 9.71 | 7.24 | 6.66 | 6.02 |
| 79 | 10.51 | 7.26 | 6.64 | 5.96 | 9.12 | 6.75 | 6.24 | 5.67 |
| 80 | 9.86 | 6.76 | 6.21 | 5.61 | 8.55 | 6.27 | 5.82 | 5.31 |
| 81 | 9.22 | 6.29 | 5.81 | 5.27 | 8.00 | 5.83 | 5.43 | 4.99 |
| 82 | 8.61 | 5.83 | 5.42 | 4.94 | 7.46 | 5.39 | 5.05 | 4.66 |
| 83 | 8.02 | 5.39 | 5.03 | 4.61 | 6.95 | 4.97 | 4.67 | 4.33 |
| 84 | 7.46 | 4.97 | 4.65 | 4.28 | 6.46 | 4.56 | 4.30 | 4.00 |
| 85 | 6.92 | 4.56 | 4.28 | 3.96 | 6.00 | 4.17 | 3.95 | 3.69 |
| 86 | 6.41 | 4.22 | 3.98 | 3.70 | 5.57 | 3.84 | 3.65 | 3.43 |
| 87 | 5.94 | 3.90 | 3.69 | 3.45 | 5.17 | 3.53 | 3.37 | 3.18 |
| 88 | 5.49 | 3.60 | 3.42 | 3.21 | 4.79 | 3.24 | 3.10 | 2.93 |
| 89 | 5.08 | 3.31 | 3.17 | 2.99 | 4.44 | 2.96 | 2.84 | 2.70 |
| 90 | 4.68 | 3.05 | 2.92 | 2.77 | 4.12 | 2.69 | 2.59 | 2.47 |
| 91 | 4.32 | 2.79 | 2.69 | 2.56 | 3.80 | 2.47 | 2.39 | 2.28 |
| 92 | 3.98 | 2.55 | 2.47 | 2.36 | 3.51 | 2.27 | 2.20 | 2.11 |
| 93 | 3.67 | 2.33 | 2.26 | 2.17 | 3.23 | 2.07 | 2.02 | 1.94 |
| 94 | 3.38 | 2.11 | 2.05 | 1.98 | 2.98 | 1.89 | 1.84 | 1.78 |
| 95 | 3.11 | 1.90 | 1.85 | 1.80 | 2.75 | 1.71 | 1.67 | 1.62 |
| 96 | 2.88 | 1.69 | 1.66 | 1.61 | 2.55 | 1.53 | 1.50 | 1.47 |
| 97 | 2.67 | 1.48 | 1.45 | 1.42 | 2.38 | 1.36 | 1.34 | 1.31 |
| 98 | 2.47 | 1.23 | 1.22 | 1.20 | 2.21 | 1.15 | 1.13 | 1.12 |
| 99 | 2.28 | 0.94 | 0.93 | 0.92 | 2.04 | 0.88 | 0.87 | 0.87 |

LE = life expectancy; QALE = quality-adjusted life expectancy

Appendix Table 3 provides QALE norms based on the same 2017-2019 life tables but using the original MVH EQ-5D-3L population norms (MVH Group 1995). The resulting QALE estimates are considerably higher than those reported in Table 2. For example, the estimated undiscounted QALE at birth for females is 71.9 QALYs or nearly 3.2 QALYs more than estimated based on the -5L population norms reported in Table 1. A similar albeit smaller gap is observed for men (69.2 vs. 68.3 QALYs).

**Table 3.** QALE by age and sex with 0%, 1.5% and 3.5% discount rates (based on MVH population norms for HRQoL (EQ-5D-3L health states and UK social value set) and 2017-2019 life tables.

| Age | Female |  |  |  | Male |  |  |  |
| --- | --- | --- | --- | --- | --- | --- | --- | --- |
|  | LE |  | QALE |  | LE |  | QALE |  |
|  |  | 0 % | 1.5 % | 3.5 % |  | 0 % | 1.5 % | 3.5 % |
| 0 | 83.33 | 71.93 | 42.39 | 25.23 | 79.67 | 69.19 | 41.42 | 24.95 |
| 1 | 82.63 | 71.25 | 42.23 | 25.23 | 79.02 | 68.54 | 41.27 | 24.96 |
| 2 | 81.65 | 70.33 | 41.92 | 25.15 | 78.04 | 67.62 | 40.94 | 24.87 |
| 3 | 80.66 | 69.39 | 41.60 | 25.06 | 77.05 | 66.69 | 40.61 | 24.77 |
| 4 | 79.67 | 68.46 | 41.27 | 24.96 | 76.06 | 65.76 | 40.27 | 24.66 |
| 5 | 78.67 | 67.52 | 40.94 | 24.86 | 75.06 | 64.82 | 39.92 | 24.55 |
| 6 | 77.68 | 66.59 | 40.60 | 24.76 | 74.07 | 63.89 | 39.57 | 24.44 |
| 7 | 76.68 | 65.66 | 40.26 | 24.66 | 73.08 | 62.95 | 39.21 | 24.33 |
| 8 | 75.69 | 64.72 | 39.91 | 24.55 | 72.08 | 62.02 | 38.85 | 24.21 |
| 9 | 74.69 | 63.78 | 39.56 | 24.44 | 71.09 | 61.08 | 38.48 | 24.08 |
| 10 | 73.70 | 62.85 | 39.20 | 24.32 | 70.09 | 60.15 | 38.11 | 23.96 |
| 11 | 72.70 | 61.91 | 38.84 | 24.20 | 69.09 | 59.21 | 37.73 | 23.82 |
| 12 | 71.71 | 60.98 | 38.47 | 24.08 | 68.10 | 58.27 | 37.34 | 23.69 |
| 13 | 70.71 | 60.04 | 38.09 | 23.95 | 67.11 | 57.34 | 36.95 | 23.55 |
| 14 | 69.72 | 59.10 | 37.71 | 23.82 | 66.12 | 56.41 | 36.56 | 23.40 |
| 15 | 68.72 | 58.17 | 37.33 | 23.68 | 65.12 | 55.47 | 36.15 | 23.25 |
| 16 | 67.73 | 57.23 | 36.94 | 23.54 | 64.13 | 54.54 | 35.75 | 23.09 |
| 17 | 66.74 | 56.30 | 36.54 | 23.39 | 63.15 | 53.62 | 35.34 | 22.93 |
| 18 | 65.75 | 55.37 | 36.14 | 23.24 | 62.17 | 52.69 | 34.93 | 22.77 |
| 19 | 64.76 | 54.44 | 35.74 | 23.09 | 61.19 | 51.77 | 34.51 | 22.60 |
| 20 | 63.78 | 53.51 | 35.33 | 22.93 | 60.22 | 50.85 | 34.09 | 22.43 |
| 21 | 62.79 | 52.58 | 34.91 | 22.76 | 59.25 | 49.94 | 33.66 | 22.25 |
| 22 | 61.80 | 51.65 | 34.48 | 22.59 | 58.28 | 49.02 | 33.23 | 22.07 |
| 23 | 60.81 | 50.72 | 34.06 | 22.41 | 57.30 | 48.11 | 32.79 | 21.88 |
| 24 | 59.82 | 49.79 | 33.62 | 22.23 | 56.33 | 47.19 | 32.34 | 21.69 |
| 25 | 58.84 | 48.87 | 33.18 | 22.04 | 55.36 | 46.27 | 31.89 | 21.48 |
| 26 | 57.85 | 47.95 | 32.74 | 21.85 | 54.39 | 45.37 | 31.44 | 21.28 |
| 27 | 56.87 | 47.03 | 32.29 | 21.66 | 53.42 | 44.46 | 30.99 | 21.08 |
| 28 | 55.88 | 46.11 | 31.84 | 21.46 | 52.45 | 43.56 | 30.52 | 20.87 |
| 29 | 54.90 | 45.20 | 31.39 | 21.26 | 51.48 | 42.65 | 30.06 | 20.65 |
| 30 | 53.92 | 44.28 | 30.92 | 21.05 | 50.51 | 41.75 | 29.58 | 20.42 |
| 31 | 52.93 | 43.37 | 30.45 | 20.83 | 49.55 | 40.85 | 29.11 | 20.19 |
| 32 | 51.95 | 42.45 | 29.98 | 20.60 | 48.59 | 39.95 | 28.62 | 19.95 |
| 33 | 50.98 | 41.54 | 29.50 | 20.37 | 47.62 | 39.05 | 28.13 | 19.70 |
| 34 | 50.00 | 40.63 | 29.01 | 20.13 | 46.66 | 38.16 | 27.63 | 19.44 |
| 35 | 49.03 | 39.72 | 28.51 | 19.88 | 45.71 | 37.26 | 27.12 | 19.18 |
| 36 | 48.05 | 38.83 | 28.03 | 19.65 | 44.75 | 36.38 | 26.63 | 18.93 |
| 37 | 47.08 | 37.94 | 27.55 | 19.41 | 43.80 | 35.51 | 26.14 | 18.67 |
| 38 | 46.11 | 37.06 | 27.06 | 19.16 | 42.85 | 34.65 | 25.64 | 18.40 |
| 39 | 45.15 | 36.18 | 26.56 | 18.90 | 41.9 | 33.78 | 25.13 | 18.12 |
| 40 | 44.18 | 35.29 | 26.05 | 18.63 | 40.95 | 32.91 | 24.61 | 17.84 |
| 41 | 43.22 | 34.41 | 25.54 | 18.36 | 40.01 | 32.04 | 24.10 | 17.55 |
| 42 | 42.26 | 33.53 | 25.03 | 18.08 | 39.07 | 31.18 | 23.57 | 17.25 |
| 43 | 41.30 | 32.66 | 24.50 | 17.79 | 38.14 | 30.33 | 23.04 | 16.94 |
| 44 | 40.34 | 31.78 | 23.97 | 17.49 | 37.21 | 29.47 | 22.50 | 16.62 |
| 45 | 39.39 | 30.91 | 23.44 | 17.18 | 36.28 | 28.62 | 21.96 | 16.29 |
| 46 | 38.45 | 30.10 | 22.96 | 16.92 | 35.36 | 27.84 | 21.49 | 16.03 |
| 47 | 37.50 | 29.30 | 22.48 | 16.66 | 34.44 | 27.06 | 21.01 | 15.76 |
| 48 | 36.56 | 28.50 | 21.99 | 16.39 | 33.53 | 26.29 | 20.52 | 15.48 |
| 49 | 35.63 | 27.70 | 21.49 | 16.12 | 32.62 | 25.52 | 20.03 | 15.20 |

LE = life expectancy; QALE = quality-adjusted life expectancy

**Table 3. (continued)**
| Age | Female |  |  |  | Male |  |  |  |
| --- | --- | --- | --- | --- | --- | --- | --- | --- |
|  | LE |  | QALE |  | LE |  | QALE |  |
|  |  | 0 % | 1.5 % | 3.5 % |  | 0 % | 1.5 % | 3.5 % |
| 50 | 34.69 | 26.90 | 20.99 | 15.83 | 31.71 | 24.76 | 19.54 | 14.91 |
| 51 | 33.77 | 26.10 | 20.49 | 15.54 | 30.81 | 24.00 | 19.05 | 14.61 |
| 52 | 32.84 | 25.31 | 19.98 | 15.24 | 29.92 | 23.24 | 18.54 | 14.30 |
| 53 | 31.92 | 24.52 | 19.47 | 14.93 | 29.03 | 22.49 | 18.04 | 13.99 |
| 54 | 31.00 | 23.74 | 18.95 | 14.61 | 28.15 | 21.74 | 17.53 | 13.67 |
| 55 | 30.09 | 22.95 | 18.42 | 14.29 | 27.27 | 20.99 | 17.02 | 13.33 |
| 56 | 29.18 | 22.21 | 17.93 | 13.99 | 26.39 | 20.30 | 16.56 | 13.06 |
| 57 | 28.28 | 21.47 | 17.44 | 13.69 | 25.53 | 19.63 | 16.10 | 12.77 |
| 58 | 27.38 | 20.74 | 16.94 | 13.38 | 24.67 | 18.96 | 15.64 | 12.49 |
| 59 | 26.49 | 20.02 | 16.44 | 13.07 | 23.82 | 18.29 | 15.18 | 12.19 |
| 60 | 25.61 | 19.29 | 15.94 | 12.74 | 22.98 | 17.63 | 14.72 | 11.90 |
| 61 | 24.73 | 18.57 | 15.43 | 12.41 | 22.14 | 16.98 | 14.25 | 11.59 |
| 62 | 23.86 | 17.86 | 14.92 | 12.08 | 21.32 | 16.34 | 13.79 | 11.29 |
| 63 | 23.00 | 17.16 | 14.41 | 11.74 | 20.51 | 15.70 | 13.33 | 10.98 |
| 64 | 22.15 | 16.46 | 13.90 | 11.38 | 19.71 | 15.07 | 12.87 | 10.66 |
| 65 | 21.3 | 15.76 | 13.38 | 11.02 | 18.91 | 14.45 | 12.41 | 10.34 |
| 66 | 20.46 | 15.10 | 12.89 | 10.69 | 18.13 | 13.84 | 11.95 | 10.02 |
| 67 | 19.62 | 14.44 | 12.40 | 10.34 | 17.36 | 13.24 | 11.49 | 9.69 |
| 68 | 18.80 | 13.79 | 11.91 | 9.99 | 16.6 | 12.64 | 11.03 | 9.36 |
| 69 | 17.98 | 13.14 | 11.41 | 9.63 | 15.86 | 12.05 | 10.57 | 9.03 |
| 70 | 17.17 | 12.50 | 10.91 | 9.27 | 15.12 | 11.47 | 10.11 | 8.69 |
| 71 | 16.38 | 11.87 | 10.41 | 8.90 | 14.38 | 10.89 | 9.65 | 8.34 |
| 72 | 15.58 | 11.24 | 9.91 | 8.52 | 13.66 | 10.32 | 9.20 | 7.99 |
| 73 | 14.81 | 10.62 | 9.41 | 8.13 | 12.96 | 9.76 | 8.74 | 7.64 |
| 74 | 14.06 | 10.02 | 8.92 | 7.75 | 12.27 | 9.22 | 8.30 | 7.29 |
| 75 | 13.31 | 9.42 | 8.43 | 7.36 | 11.6 | 8.69 | 7.85 | 6.94 |
| 76 | 12.58 | 8.90 | 8.00 | 7.03 | 10.95 | 8.20 | 7.45 | 6.62 |
| 77 | 11.87 | 8.40 | 7.59 | 6.71 | 10.32 | 7.73 | 7.05 | 6.31 |
| 78 | 11.18 | 7.90 | 7.18 | 6.39 | 9.71 | 7.27 | 6.67 | 5.99 |
| 79 | 10.51 | 7.43 | 6.78 | 6.07 | 9.12 | 6.83 | 6.29 | 5.68 |
| 80 | 9.86 | 6.96 | 6.39 | 5.75 | 8.55 | 6.39 | 5.92 | 5.38 |
| 81 | 9.22 | 6.51 | 6.01 | 5.43 | 8.00 | 5.98 | 5.56 | 5.08 |
| 82 | 8.61 | 6.07 | 5.63 | 5.12 | 7.46 | 5.58 | 5.21 | 4.79 |
| 83 | 8.02 | 5.65 | 5.26 | 4.82 | 6.95 | 5.19 | 4.87 | 4.49 |
| 84 | 7.46 | 5.25 | 4.91 | 4.52 | 6.46 | 4.82 | 4.54 | 4.21 |
| 85 | 6.92 | 4.87 | 4.57 | 4.23 | 6.00 | 4.48 | 4.23 | 3.95 |
| 86 | 6.41 | 4.50 | 4.25 | 3.95 | 5.57 | 4.15 | 3.94 | 3.69 |
| 87 | 5.94 | 4.16 | 3.94 | 3.69 | 5.17 | 3.85 | 3.66 | 3.44 |
| 88 | 5.49 | 3.84 | 3.65 | 3.43 | 4.79 | 3.56 | 3.40 | 3.21 |
| 89 | 5.08 | 3.54 | 3.38 | 3.19 | 4.44 | 3.29 | 3.16 | 2.99 |
| 90 | 4.68 | 3.25 | 3.12 | 2.95 | 4.12 | 3.05 | 2.93 | 2.79 |
| 91 | 4.32 | 2.98 | 2.87 | 2.73 | 3.8 | 2.80 | 2.70 | 2.58 |
| 92 | 3.98 | 2.73 | 2.63 | 2.52 | 3.51 | 2.57 | 2.49 | 2.39 |
| 93 | 3.67 | 2.48 | 2.41 | 2.31 | 3.23 | 2.35 | 2.28 | 2.20 |
| 94 | 3.38 | 2.25 | 2.19 | 2.11 | 2.98 | 2.13 | 2.08 | 2.01 |
| 95 | 3.11 | 2.02 | 1.98 | 1.92 | 2.75 | 1.93 | 1.89 | 1.84 |
| 96 | 2.88 | 1.80 | 1.77 | 1.72 | 2.55 | 1.73 | 1.70 | 1.66 |
| 97 | 2.67 | 1.58 | 1.55 | 1.52 | 2.38 | 1.54 | 1.51 | 1.49 |
| 98 | 2.47 | 1.31 | 1.30 | 1.28 | 2.21 | 1.30 | 1.28 | 1.27 |
| 99 | 2.28 | 1.00 | 0.99 | 0.99 | 2.04 | 0.99 | 0.99 | 0.98 |

LE = life expectancy; QALE = quality-adjusted life expectancy

## Discussion

NICE is proposing to introduce severity-of-disease modifiers that would assign greater value to QALY gains for patients with greater absolute or relative expected shortfall in QALE under the current standard of care. This short note provides updated QALE population norms based on the EQ-5D-5L instrument that serve to establish the benchmark against which shortfalls can be assessed. These population norms combine official, full population life tables with HRQoL data obtained from a large, representative sample of the English population. Additional data tables and figures are made available through an interactive web application, available at: https://r4scharr.shinyapps.io/shortfall/.

There are three main limitations to our study: First, both life tables and HRQoL data reflect the health of current populations and, as a result, our estimates should be interpreted as period QALEs. Medical and societal progress are likely to change both LE and HRQoL for future cohorts of patients, thus creating a need for regular updates of these QALE population norms. Second, approximately 10% of the participants in the HSE did not report their EQ-5D-5L health profile and were therefore not included in the study. This might have introduced selection bias in our estimates of average HRQoL by age and sex. However, previous work by Love-Koh et al. (2015) found that imputing missing HRQoL data changed QALE estimates by less than 0.01 QALYs. We therefore believe that missing data are unlikely to introduce significant bias. Finally, our analysis is based on EQ-5D-5L data being mapped to and valued using the EQ-5D-3L value set (i.e. cross-walking), which is consistent with NICE’s current reference case. A new valuation study for the EQ-5D-5L is underway (EuroQol 2020), which will provide health state valuations without the need for crosswalks and associated loss of information. Once published, we plan to update the interactive website to give stakeholders access to QALE estimates based on this new value set.

## Conclusion

This study provides new QALE population norms for England. These norms serve as an input for the calculation of absolute and relative QALY shortfalls to inform health technology assessment with severity of condition adjustment as currently considered for implementation in England.

## Data Availability

All the data set that were used are publicly available. The source of the analysis and the web application is available at: https://github.com/bitowaqr/shortfall

https://r4scharr.shinyapps.io/shortfall/

## Funding and acknowledgements

This work was supported by the EuroQol Foundation (project number: 123-2020RA), the Wellcome Trust Doctoral Training Centre in Public Health Economics and Decision Science (108903/Z/19/Z), and the University of Sheffield through a PhD scholarship. We are grateful to Matthijs Versteegh for providing comments on an earlier version of this manuscript. The usual disclaimer applies.

## References

Arneberg, F. (2012). Measuring the level of severity in pharmacoeconomic analyses: an empirical approach. Reprosentralen, University of Oslo. url: https://www.duo.uio.no/bitstream/handle/10852/30279/Masterx-xArneberg.pdf?sequence=1 (last accessed 10/12/2021)

Catchpole, P., Barrett, V. (2020). Keeping Pace with Pharmaceutical Innovation: The Importance of the NICE Methods Review. PharmacoEconomics 38, 901–903.

Chiang, C. L. (1984). Life table and its applications. Florida: Krieger.

EuroQol (2020). New UK EQ-5D-5L Valuation Study. url: https://euroqol.org/eq-5d-instruments/eq-5d-5l-about/valuation-standard-value-sets/new-uk-eq-5d-5l-valuation-study_blog/ (last accessed 10/12/2021)

Heijink, R., Van Baal, P., Oppe, M., Koolman, X., Westert, G. (2011). Decomposing cross-country differences in quality adjusted life expectancy: the impact of value sets. Population health metrics, 9(1), 1–11.

Herdman, M., Gudex, C., Lloyd, A., Janssen, M. F., Kind, P., Parkin, D., … Badia, X. (2011). Development and preliminary testing of the new five-level version of EQ-5D (EQ-5D-5L). Quality of life research, 20(10), 1727–1736.

Kind, P., Hardman, G., Macran, S. (1999). UK population norms for EQ-5D (No. 172chedp). Centre for Health Economics, University of York.

Love-Koh, J., Asaria, M., Cookson, R., Griffin, S. (2015). The social distribution of health: estimating quality-adjusted life expectancy in England. Value in health, 18(5), 655–662.

MVH Group (1995). The measurement and valuation of health: Final report on the modelling of valuation tariffs. Centre for Health Economics, University of York. url: https://www.york.ac.uk/media/che/documents/reports/MVHFinalReport.pdf (last accessed 10/12/2021)

NatCen (2019). Health Survey for England, 2017 [data collection]. 2nd Edition. University College London Department of Epidemiology and Public Health National Centre for Social Research. UK Data Service. SN: 8488, 2019.

NatCen (2020). Health Survey for England, 2018 [data collection]. 2nd Edition. University College London Department of Epidemiology and Public Health National Centre for Social Research. UK Data Service. SN: 8649.

National Institute for Health and Clinical Excellence (2009). Appraising life-extending, end of life treatments. url: https://www.nice.org.uk/guidance/GID-TAG387/documents/appraising-life-extending-end-of-life-treatments-paper2 (last accessed 10/12/2021)

National Institute for Health and Care Excellence (2021). Methods, processes and topic selection for health technology evaluation: proposals for change. url: https://www.nice.org.uk/about/what-we-do/our-programmes/nice-guidance/chte-methods-and-processes-consultation (last accessed 10/12/2021)

Office for National Statistics (2021). National Life Tables, England, 1980-1982 to 2017-2019. Url: https://www.ons.gov.uk/peoplepopulationandcommunity/birthsdeathsandmarriages/lifeexpectancies/datasets/nationallifetablesenglandreferencetables (last accessed 10/12/2021)

Ottersen T, Førde R, Kakad M, Kjellevold A, Melberg H, Moen A, Ringard A, Norheim O. (2016). A new proposal for priority setting in Norway: open and fair. Health Policy. 120, 246–251

Palmer, A. J., Campbell, J. A., de Graaff, B., Devlin, N., Ahmad, H., Clarke, P. M., … Si, L. (2021). Population norms for quality adjusted life years for the United States of America, China, the United Kingdom and Australia. Health Economics. 30:1950–1977

Reckers-Droog, V. T., Van Exel, N. J. A., Brouwer, W. B. F. (2018). Looking back and moving forward: on the application of proportional shortfall in healthcare priority setting in the Netherlands. Health policy, 122(6), 621–629.

Statens legemiddelverk (2020). Guidelines for the submission of documentation for single technology assessment (STA) of pharmaceuticals. url: https://legemiddelverket.no/Documents/English/Public\%20funding\%20and\%20pricing/Documentation\%20for\%20STA/Guidelines\%2020.05.2020.pdf (last accessed 10/12/2021)

Stolk EA, van Donselaar G, Brouwer WB, Busschbach JJ. (2004). Reconciliation of economic concerns and health policy: illustration of an equity adjustment procedure using proportional shortfall. Pharmacoeconomics. 22(17):1097–107.

Sullivan, D. F. (1971). A single index of mortality and morbidity. HSMHA health reports, 86(4), 347.

Van Hout, B., Janssen, M. F., Feng, Y. S., Kohlmann, T., Busschbach, J., Golicki, D., … Pickard, A. S. (2012). Interim scoring for the EQ-5D-5L: mapping the EQ-5D-5L to EQ-5D-3L value sets. Value in health, 15(5), 708–715.

Versteegh, M. M., Ramos, I. C., Buyukkaramikli, N. C., Ansaripour, A., Reckers-Droog, V. T., Brouwer, W. B. (2019). Severity-adjusted probability of being cost effective. Pharmacoeconomics, 37(9), 1155–1163. iDBC tool: https://imta.shinyapps.io/iDBC/

Zorginstituut Nederland. (2015). Cost-effectiveness in practice. url: https://www.zorginstituutnederland.nl/publicaties/rapport/2015/06/26/kosteneffectiviteit-in-de-praktijk. (last accessed 10/12/2021)

